# Increased Risk of Pulmonary Embolism Following SARS-CoV-2 Activity in Ontario, Canada

**DOI:** 10.64898/2026.03.27.26349516

**Authors:** Clara Eunyoung Lee, Natalie J. Wilson, David N. Fisman

## Abstract

**Background:** SARS-CoV-2 infection is an established prothrombotic trigger, yet the population-level temporal relationship between circulating viral activity and pulmonary embolism (PE) remains poorly characterized. We aimed to evaluate the short-term association between respiratory viral activity and PE hospitalizations, accounting for specific temporal lags.

**Methods:** We conducted a population-level time-series analysis of incident PE hospitalizations in Ontario, Canada, from 2011 to 2024. Using distributed lag non-linear models, we assessed the association between standardized weekly activity levels of SARS-CoV-2, influenza A/B, and respiratory syncytial virus (RSV) and PE risk over a 5-week lag period. Relative risks (RR) per standard deviation (SD) increase in viral activity were estimated via negative binomial regression using cross-basis terms to account for both exposure-response and lag-response non-linearities. Models were adjusted for Fourier seasonal terms and secular trends.

**Findings:** Among 70,670 incident PE cases identified between 2011 and 2024, SARS-CoV-2 activity demonstrated a significant temporal association with PE. A cumulative RR increase of 20% per SD in SARS-CoV-2 activity was observed over the five weeks following exposure (RR 1.20; 95% CI 1.05–1.37). The risk followed a distinct delay trajectory: weekly cumulative RRs peaked at week 3 (RR 1.21; 95% CI 1.01–1.45). For the 2020-2024 period, influenza A also showed an association peaking at week 3 without statistical significance (RR 1.17; 95% CI 0.95–1.45).

**Interpretation:** Increased population-level SARS-CoV-2 activity is associated with a heightened risk of PE, peaking at approximately the third week. This delayed peak suggests a protracted thrombo-inflammatory window, likely driven by sustained endothelial injury. These findings highlight the vascular burden of COVID-19 and suggest that infection prevention measures, including vaccination, may provide significant downstream protection against thromboembolic disease.

## INTRODUCTION

SARS-CoV-2 infection is linked to an increased risk of venous thromboembolism^1,2^ and is associated with high risk of mortality. Early histopathological evidence from autopsies of individuals who died with SARS-CoV-2 infection revealed deep venous thrombosis in 58% of decedents, with pulmonary embolism (PE) identified as the primary cause of death one-third of cases.^3^ Subsequent large-scale epidemiological studies have confirmed this association, with one Swedish cohort study reporting a 33-fold increase in PE risk in the 30 days following infection.^4^

The pathophysiology of COVID-19-induced thrombosis is multifactorial, involving direct endothelial injury, neutrophil and platelet activation, and a “cytokine storm” characterized by the overproduction of thrombogenic cytokines.^5^ Because these inflammatory processes evolve progressively, PE often manifests after a significant clinical delay. This delay may make it challenging to evaluate linkages between SARS-CoV-2 infection and PE, particularly when many individuals do not undergo virological testing, and mild or asymptomatic infections remain undiagnosed.

While individual-level studies support the association between SARS-CoV-2 infection and PE, the timing remains less well characterized at the population level. Much research has focused on the acute 90-day post-infection window without delineating daily or weekly temporal lag patterns of onset.^6^ Furthermore, reliance on confirmed cases introduces significant selection bias toward hospitalized or highly symptomatic patients, and those with comorbidities (such as cancer) that may increase both COVID-19 severity and PE risk.

In this study, we sought to address these gaps by treating publicly reported respiratory viral surveillance data as an environmental exposure, and as a proxy for individual risk, thus avoiding pitfalls associated with selection bias in highly tested populations. By applying distributed lag non-linear models (DLNMs) to provincial administrative health records, we characterized the short-term temporal relationship between population-level viral activity and subsequent PE hospitalizations, and provide a clear estimate of the burden of pulmonary thromboembolic disease attributable to SARS-CoV-2 during the pandemic.

## METHODS

### Data Sources and Study Population Construction

The study area comprised the Greater Toronto Area (GTA) and adjacent regions in Ontario, Canada.^7^ As Canada’s most populous metropolitan region, the GTA had a population of 6,202,225 in 2021 and encompasses a total area of 5,902.75 km^2^. ^8^

Hospitalization data for PE were obtained from the Canadian Institutes of Health Information (CIHI) Discharge Abstract Database. Under Canada’s universal healthcare system, CIHI maintains a comprehensive national repository of all acute care hospitalizations, ensuring high data accessibility and completeness.^9^ We restricted our analysis to inpatient data to ensure the inclusion of clinically significant PE cases requiring hospitalization, consistent with established methodologies.^10,11^

PE cases were identified using the International Classification of Diseases, 10^th^ Revision (ICD-10) codes I26.0 and I26.9 from all 25 available diagnosis fields. To ensure each patient was represented only once, we restricted the cohort to incident PE events by selecting the index hospitalization date for each individual. Subsequent readmissions or repeat diagnosis codes for the same individual were excluded to prevent the inflation of risk estimates due to recurrent events. Anonymized individual-level records were aggregated into a weekly time series covering the period from 2011to 2024.

### Respiratory Viral Surveillance and Activity Characterization

Daily surveillance data for SARS-CoV-2, influenza A and B, and respiratory syncytial virus (RSV) were sourced from publicly available data from Public Health Ontario and the FluWatch program. Viral activity records were extracted for the full study period (2011- 2024) for influenza and RSV, while SARS-CoV-2 data were collected from its emergence in 2020 through 2024.

To facilitate comparison across different viruses and evolving surveillance systems, exposure variables were standardized by their respective study-period standard deviations (SD). This scaling approach allowed for a uniform analysis of viral activity regardless of the original measurement metric (e.g., percent positivity or absolute case counts). By preserving the natural zero-point of the surveillance data, this method ensures that the baseline for the relative risk (RR=1.0) corresponds to absent viral circulation, while enabling the estimation of risk per unit SD increase in population-level viral activity.

### Statistical Modeling and Lag-Response Characterization

We utilized a distributed lag non-linear modeling (DLNM) framework integrated within negative binomial regression models to characterize the exposure-lag-response relationship between viral activity and PE hospitalizations. The negative binomial approach was selected to account for overdispersion in weekly PE counts.

For each virus, a cross-basis matrix modeled the association across a 5-week lag window, consistent with evidence that thromboembolic risk of venous thromboembolism is most pronounced within the first month post-infection. ^12^ The lag-response dimension was defined using a natural cubic spline with three degrees of freedom.

The primary analysis utilized a multi-pathogen model for influenza A, influenza B, and RSV, while SARS-CoV-2 was modeled independently to accommodate its distinct period of circulation. All models were adjusted for seasonality using Fourier terms (first- and second-order sine and cosine functions) and for long-term secular trends using a linear term for the calendar year. Model selection was determined by the lowest Akaike Information Criterion (AIC). Relative risks (RRs) were estimated per one SD increase in viral activity. We reported both RRs for discrete weekly lags and the total cumulative RR over the observation period.

### Sensitivity Analyses

To account for shifts in viral transmission dynamics and healthcare utilization during the COVID-19 pandemic, we conducted sensitivity analyses stratified by era. For the pre-pandemic era (2011–2019), we utilized a three-pathogen model (influenza A, influenza B, and RSV). For the pandemic era (2020–2024), a multi-pathogen model was developed that simultaneously incorporated cross-basis terms for SARS-CoV-2, influenza A, and RSV. Influenza B was excluded from the pandemic-era model due to its minimal circulation during this period.

All data cleaning and regression analyses were conducted within the Secure Access Environment of CIHI using R version 4.2.3, with distributed lag models constructed using the *dlnm* package. The study was approved by the Research Ethics Board, University of Toronto (Protocol # 41690). Informed consent was waived as the study utilized anonymized, pre-collected administrative datasets.

## FINDINGS

### Descriptive Epidemiology of Study Population and Viral Activity Trends

Among a total of 85,803 hospitalization for PE, 70,670 incident admissions were included in the final analysis. Throughout the study period, weekly PE cases exhibited a steady increase, followed by a sharp escalation after 2020 (**Figure 1**). This pandemic-era surge was characterized by two distinct peaks, after which case count returned to a trajectory consistent with pre-2020 trends.

**Figure 1.**
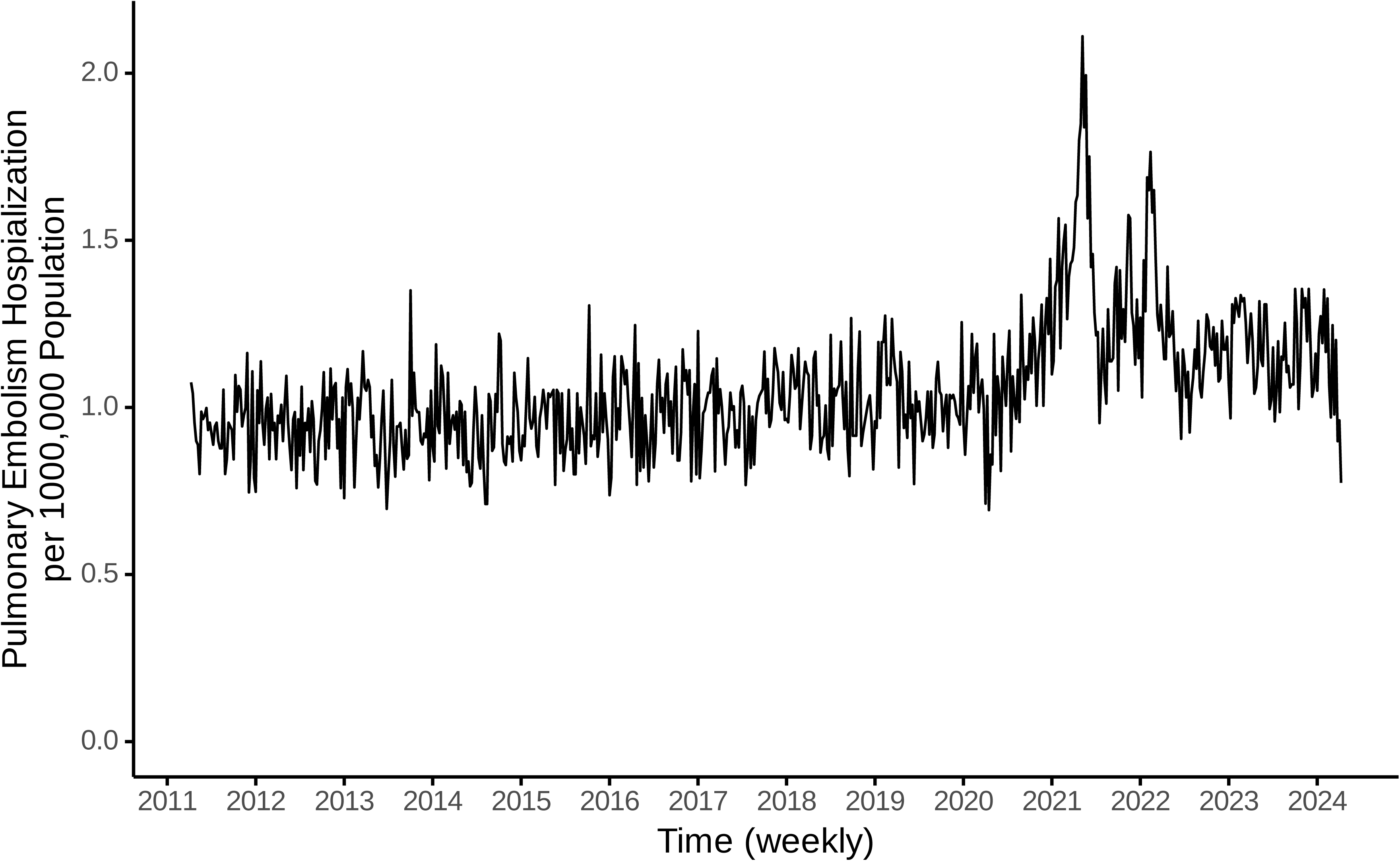
Temporal Trends in Respiratory Viral Activity and Pulmonary Embolism Hospitalizations (2011- 2024), Ontario, Canada. (A) Weekly frequency of incident admissions for pulmonary embolism (PE), totaling 70,670 cases, expressed per 100,000 population. A steady secular increase in baseline cases likely reflects regional population growth. Post 2020, PE hospitalizations rose sharply with two distinct peaks. (B) Surveillance activity of SARS-CoV-2, influenza A, influenza B, and Respiratory Syncytial virus (RSV). Pre-2020, influenza and RSV showed clear winter seasonality. This pattern was disrupted in 2020-2021 by multiple SARS-CoV-2 epidemic waves and a marked decline in other respiratory viruses. Since late 2021, RSV and influenza A have resumed seasonal periodicity, while SARS-CoV-2 transitioned to an endemic pattern with sustained activity through 2024.

Between 2011 and 2019, respiratory viral activity followed a predictable seasonal pattern with influenza A, influenza B, and RSV predominating during the winter months. Following the emergence of COVID-19 in 2020, activity of these three viruses decreased markedly, while SARS-CoV-2 exhibited multiple epidemic waves through 2024. Starting in late 2021, RSV activity rebounded and resumed its typical winter predominance. Influenza A activity similarly increased beginning in 2022, whereas influenza B demonstrated minimal circulation throughout the pandemic period.

### Temporal Association Between Respiratory Viral Activity and Pulmonary Embolism

SARS-CoV-2 was associated with an increased cumulative RR of 1.20 (95% CI 1.05-1.37) over the study period (Table 1). The highest risk was observed in week 3 (RR 1.21; 95% CI 1.01-1.45). The other three respiratory viruses did not show significant cumulative association with PE (influenza A cumulative RR 0.91; 95% CI 0.85-0.97, influenza B cumulative RR 0.95; 95% CI 0.88-1.02, and RSV cumulative RR 0.95; 95% CI 0.89-1.02).

**Table 1.** Interval-Specific Relative Risks of Pulmonary Embolism Hospitalization Following.

| Time Window | Influenza A (RR, 95% CI) | Influenza B (RR, 95% CI) | RSV (RR, 95% CI) | SARS-CoV-2* (RR, 95% CI) |
| --- | --- | --- | --- | --- |
| **Week 1** | 0.96(0.90–1.03) | 1.03(0.96–1.10) | 0.94(0.88–1.00) | 0.91(0.75–1.10) |
| **Week 2** | 0.99(0.95–1.02) | 0.97(0.94–1.00) | 0.96(0.93–1.00) | **1.11(1.02–1.21)** |
| **Week 3** | 1.00(0.93–1.07) | 0.95(0.89–1.01) | 0.99(0.93–1.05) | **1.21(1.01–1.45)** |
| **Week 4** | 0.99(0.96–1.02) | 0.97(0.94–1.00) | 1.02(0.98–1.05) | **1.10(1.02–1.19)** |
| **Week 5** | 0.97(0.90–1.04) | 1.03(0.96–1.09) | 1.04(0.97–1.12) | 0.90(0.74–1.09) |
| **Cumulative RR (95% CI)** | 0.91(0.85–0.97) | 0.95(0.88–1.02) | 0.95(0.89–1.02) | **1.20(1.05–1.37)** |

SARS-CoV-2, severe acute respiratory syndrome coronavirus 2; RSV, respiratory syncytial virus;
RR, relative risk; CI, confidence interval
\* Study duration is 2020-2024 for SARS-CoV-2.

In weekly lag models, SARS-CoV-2 activity demonstrated a temporal association with PE (Figure 2). Weekly cumulative RRs increased from week 1 (RR 0.91; 95% CI 0.75–1.10) to week 2 (RR 1.11; 95% CI 1.02–1.21), peaked at week 3 (RR 1.21; 95% CI 1.01–1.45) and declined at week 4 (RR 1.10; 95% CI 1.02–1.19) (Table 1). RSV showed an upward trend of weekly RR, reaching its highest estimate at week 5, though this did reach statistical significance (RR 1.04; 95%CI 0.97–1.12). Influenza A exhibited a unimodal trend, characterized by an initial increase in risk followed by a decline, although these shifts also remained statistically non-significant. Influenza B did not demonstrate a significant temporal association with PE in the weekly lag models.

**Figure 2.**
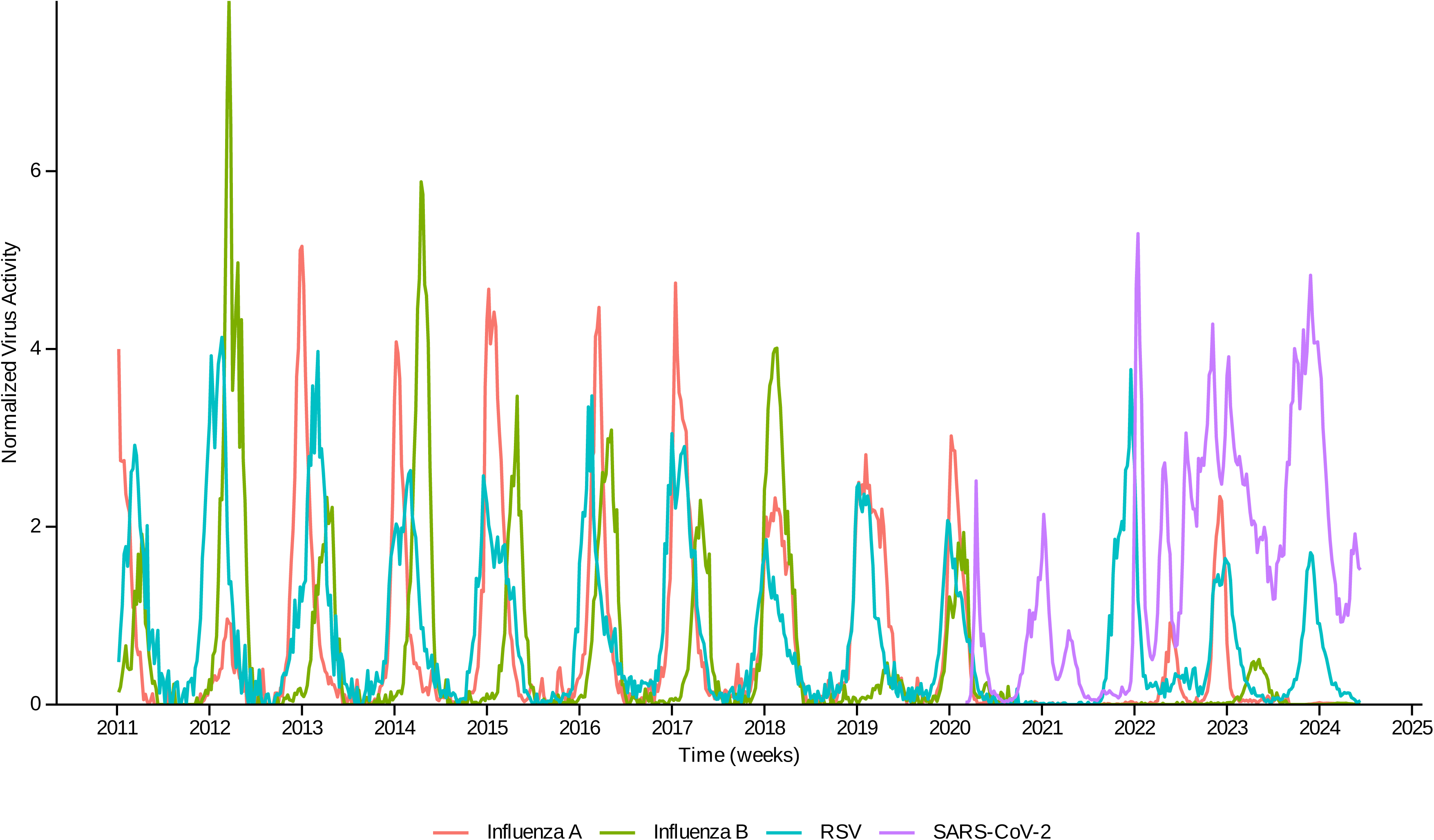
Temporal Association Between Viral Activity and Pulmonary Embolism Risk by Weekly Lag. Relative Risks (RRs) are presented for Influenza A/B and RSV (2011-2024) and SARS-CoV-2 (2020-2024). SARS-CoV-2 demonstrated a distinct temporal association, with risk peaking at Week 3 before declining by Week 4. Error bars represent 95% confidence intervals. The dashed horizontal line indicates the null effect (RR = 1.0). Points and error bars are staggered horizontally to enhance visual clarity.

The three-dimensional risk surface analysis revealed a distinct, non-linear exposure-response relationship between SARS -CoV-2 activity and PE risk. Specifically, SARS-CoV-2 exhibited a significant elevation in PE risk at a temporal lag of three weeks, with the magnitude of the relative risk increasing progressively alongside higher exposure intensity (Figure 3, Supplementary Figure 1). In contrast, the risk surfaces for influenza A, influenza B, and RSV demonstrated only subtle or negligible increase in PE risk across the range of exposure levels.

**Figure 3.**
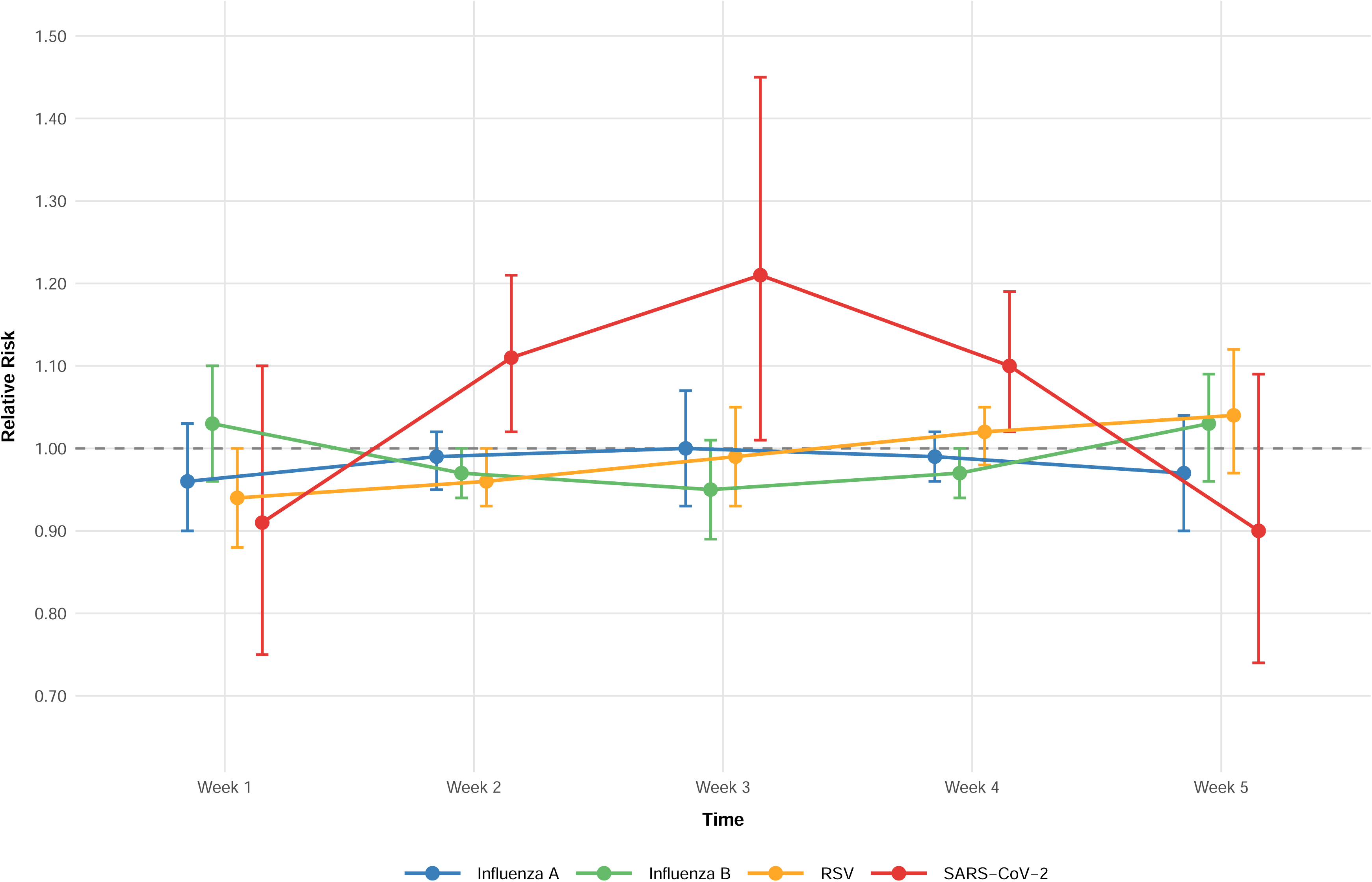
Three-dimensional Risk Surface for Pulmonary Embolism (PE) Hospitalization in Relation to Viral Exposure Intensity and Temporal Lags. The surface represents the relative risk (RR) of PE across standard deviation (SD) increases of: (A) SARS-CoV-2, (B) influenza A, (C) influenza B, and (D) Respiratory Syncytial virus (RSV). The refence for RR (1.0) is set at zero viral activity. For SARS-CoV-2, the visualization demonstrates a non-linear increase in PE risk that peaks at third week, with the magnitude of risk rising progressively with higher exposure intensity. In contrast, influenza A/B, and RSV show subtle or negligible increase in risk across exposure levels, with effect sizes consistently remaining below 1.10.

For these viruses, the effect sizes remain consistently low, with RR values remaining below 1.10 regardless of exposure intensity.

### Comparative Analysis of Pre-pandemic and Pandemic Eras

When the analysis was restricted to 2020–2024, influenza A showed a unimodal association with PE, reaching a non-significant peak at weeks 3 (RR 1.17; 95% CI 0.95–1.45; Table 2). SARS-CoV-2 maintained a concave risk profile but with a lower peak RR occurring at week 2 (RR 1.05; 95% CI 1.00–1.09), while RSV showed no significant association.

**Table 2.**
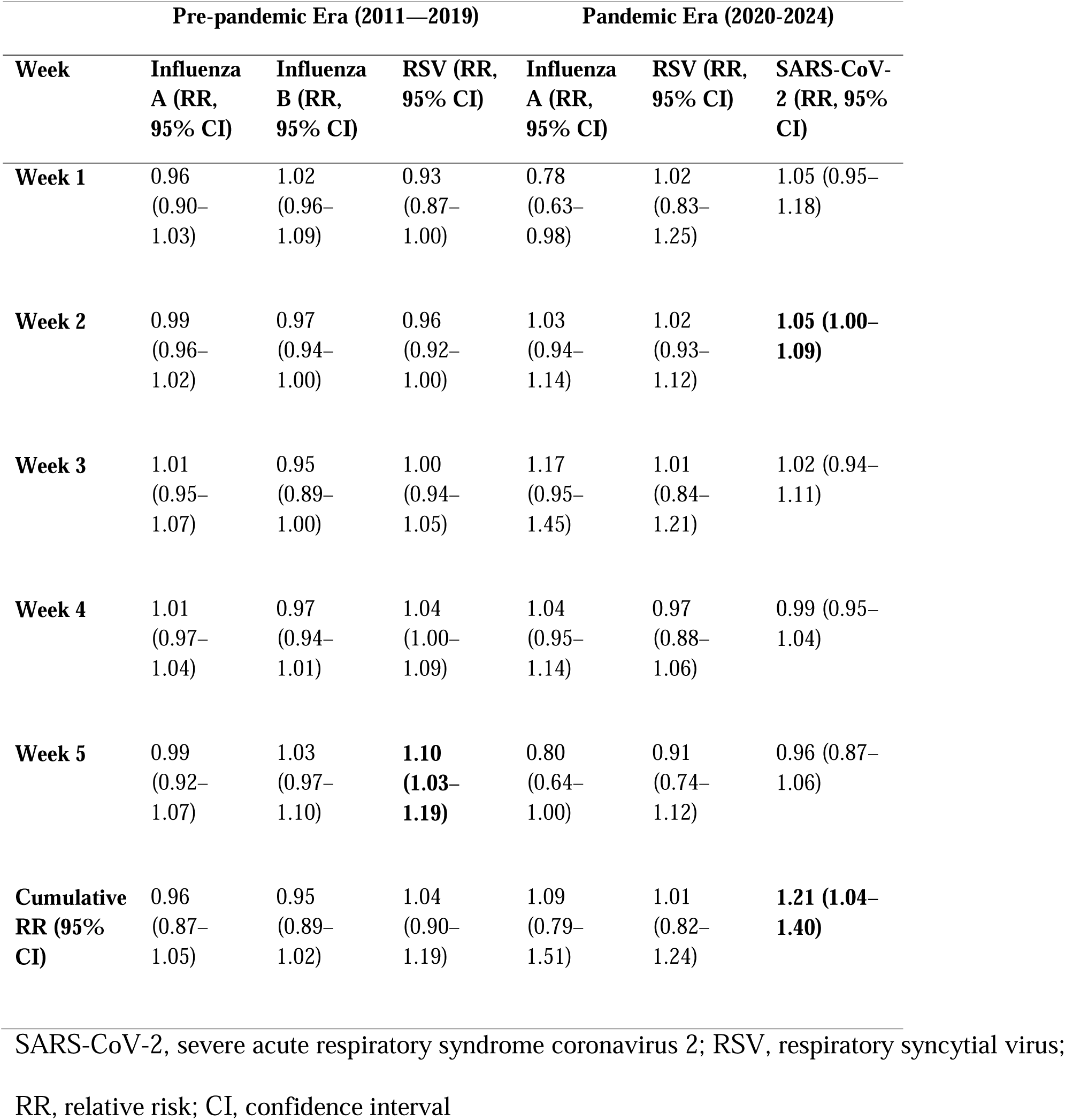
Interval-Specific Relative Risks of Pulmonary Embolism Hospitalization: A Comparison of Pre-pandemic (2011—2019) and Pandemic (2020-2024) Eras.

| Week | Pre-pandemic Era (2011—2019) |  |  | Pandemic Era (2020-2024) |  |  |
| --- | --- | --- | --- | --- | --- | --- |
|  | Influenza A (RR, 95% CI) | Influenza B (RR, 95% CI) | RSV (RR, 95% CI) | Influenza A (RR, 95% CI) | RSV (RR, 95% CI) | SARS-CoV-2 (RR, 95% CI) |
| **Week 1** | 0.96<br>(0.90–1.03) | 1.02<br>(0.96–1.09) | 0.93<br>(0.87–1.00) | 0.78<br>(0.63–0.98) | 1.02<br>(0.83–1.25) | 1.05 (0.95–1.18) |
| **Week 2** | 0.99<br>(0.96–1.02) | 0.97<br>(0.94–1.00) | 0.96<br>(0.92–1.00) | 1.03<br>(0.94–1.14) | 1.02<br>(0.93–1.12) | **1.05 (1.00–1.09)** |
| **Week 3** | 1.01<br>(0.95–1.07) | 0.95<br>(0.89–1.00) | 1.00<br>(0.94–1.05) | 1.17<br>(0.95–1.45) | 1.01<br>(0.84–1.21) | 1.02 (0.94–1.11) |
| **Week 4** | 1.01<br>(0.97–1.04) | 0.97<br>(0.94–1.01) | 1.04<br>(1.00–1.09) | 1.04<br>(0.95–1.14) | 0.97<br>(0.88–1.06) | 0.99 (0.95–1.04) |
| **Week 5** | 0.99<br>(0.92–1.07) | 1.03<br>(0.97–1.10) | **1.10**<br>**(1.03–1.19)** | 0.80<br>(0.64–1.00) | 0.91<br>(0.74–1.12) | 0.96 (0.87–1.06) |
| **Cumulative RR (95% CI)** | 0.96<br>(0.87–1.05) | 0.95<br>(0.89–1.02) | 1.04<br>(0.90–1.19) | 1.09<br>(0.79–1.51) | 1.01<br>(0.82–1.24) | **1.21 (1.04–1.40)** |

SARS-CoV-2, severe acute respiratory syndrome coronavirus 2; RSV, respiratory syncytial virus;
RR, relative risk; CI, confidence interval

Regarding cumulative risk over the 5 weeks, only SARS-CoV-2 demonstrated a statistically significant relationship with PE (cumulative RR 1.21; 95% CI 1.04–1.40). In contrast, influenza A (cumulative RR 1.09; 95% CI 0.79–1.51) and RSV (cumulative RR 1.01; 95% CI 0.82–1.24) did not reach significance.

## DISCUSSION

This population-level ecological study identifies a robust temporal association between SARS-CoV-2 activity and PE hospitalizations. Utilizing the DLNM framework, we demonstrated that the highest risk intensity occurs at week 3, a finding that aligns closely with individual-level clinical cohorts. For instance, a Swedish nationwide study reported that the PE incidence rate peaked between days 8-14 post infection (IRR 46.40; 95% CI 40.61-53.02).^4^ Similarly, a large-scale study in England and Wales found that adjusted hazard ratios for venous thromboembolism were highest in the first week (33.2; 95% CI, 31.3–35.2) but remained elevated for up to 49 weeks.^13^ Furthermore, deep vein thrombosis risk in COVID-19 outpatients is greater within the first 30 days than thereafter.^12^

The discrepancy between effect magnitude between these individual-level studies and our finding (e.g. an IRR of ∼46 vs. our RR of 1.2–1.8) likely reflects the inherent nature of our population-level ecological approach. While clinical cohort track confirmed infections, our results capture the broader community burden. By considering the population-wide attack rate of a respiratory virus, a high individual-level risk is mathematically distributed across both infected and uninfected individuals in the general population. This suggests that surges in viral circulation elevate the absolute thromboembolic risk at a population scale.

The observed three-week lag likely reflects the complex biological transition from acute infection to a subacute thrombo-inflammatory state. Initially, viral replication triggers the apoptosis of pneumocytes and endothelial cells which in turn activate platelets, induce pro-coagulant factors, and heightens systemic inflammation.^14^ While primary viral replication typically tapers within one week, this subsequent inflammatory phase is more closely correlated with clinical deterioration, pulmonary microangiopathy, and the formation of inflammatory microthrombi.^15^ Our findings suggest that the peak of this “thrombo-inflammatory cascade” manifests clinically as PE hospitalizations approximately at the third week after the initial viral surge at the population-level.

Our results confirm that the prothrombotic effect of SARS-CoV-2 is more pronounced than that of influenza A. This aligns with previous literature reporting that the adjusted hazard ratio for venous thromboembolism in COVID-19 relative to those with influenza ranges from 1.60 to 1.89, with another evidence suggesting nearly 2.5 times higher odds for PE.^6,16^

In our results, influenza A demonstrated a concave RR trend peaking at week 3, although this did not reach formal statistical significance. Due to the shared seasonal patterns of influenza A, influenza B, and RSV, isolating their individual effects remains challenging. However, a subtle shift in the peak RR for SARS-CoV-2 was observed following adjustment for these pathogens. While the primary analysis identified a peak association at week 3, this temporal window narrowed to week 2 in the multi-pathogen model. This suggests that after accounting for the overlapping seasonality effects of influenza A and RSV, the independent prothrombotic impact of SARS-CoV-2 may be more acute than clinically observed. By isolating the SARS-CoV-2 signal, it is possible that the model reduced ‘lag contamination’ from other respiratory pathogens that circulate concurrently but may possess different thrombo-inflammatory kinetics.

These findings highlight a significant population burden of SARS-CoV-2 beyond its primary respiratory manifestations. Because mRNA vaccines demonstrate robust efficacy in preventing both clinical disease and transmission, vaccination likely provides secondary protection against downstream complications such as thromboembolic disease.^17,18^ The long-term application of COVID-19 vaccination may yield broader cardiovascular benefits by mitigating infection-mediated vascular damage.

A major strength of this study is its unprecedented scale, involving over 70,000 PE cases across provincial administrative data covering 25% of the Canadian population. By treating viral activity as an environmental factor, we bypassed individual-level testing biases while capturing precise temporal patterns. However, we acknowledge that surveillance data serves as a proxy and likely underestimated the true incidence of infection, particularly as testing behavior shifted over the study period.

In conclusion, increased SARS-CoV-2 activity is a significant predictor of PE hospitalizations, characterized by a distinct temporal lag peaking in the third week. This delayed trajectory indicates that the thrombo-inflammatory process persists well beyond the acute phase of infection. Quantifying the precise magnitude and the timing of these effects provides a framework for better estimating the total SARS-CoV-2 attributable burden and underscores the importance of vaccination as a vital tool for preventing immune-mediated complications.

## Data Availability

Code available at DOI: 10.5281/zenodo.19239358. Individual-
level administrative health data were accessed within the Canadian Institute for Health
Information Secure Access Environment. Respiratory virus surveillance data are publicly
available through Public Health Agency of Canada.

## Code and data availability

Code available at DOI: 10.5281/zenodo.19239358. Individual-level administrative health data were accessed within the Canadian Institute for Health Information Secure Access Environment. Respiratory virus surveillance data are publicly available through Public Health Agency of Canada.

## ACKNOWLEDGEMENTS

Parts of this material are based on data and information provided by the Canadian Institute for Health Information (CIHI). However, the analyses, conclusions, opinions and statements expressed herein are those of the author and not necessarily those of the CIHI.

## Declaration of generative AI and AI-assisted technologies in the manuscript preparation process

During the preparation of this work the authors used Claude (Anthropic) for statistical code review and editorial revision. After using these tools, the authors reviewed and edited the content as needed and take full responsibility for the content of the published article.

---

**Supplementary Figure 1.**
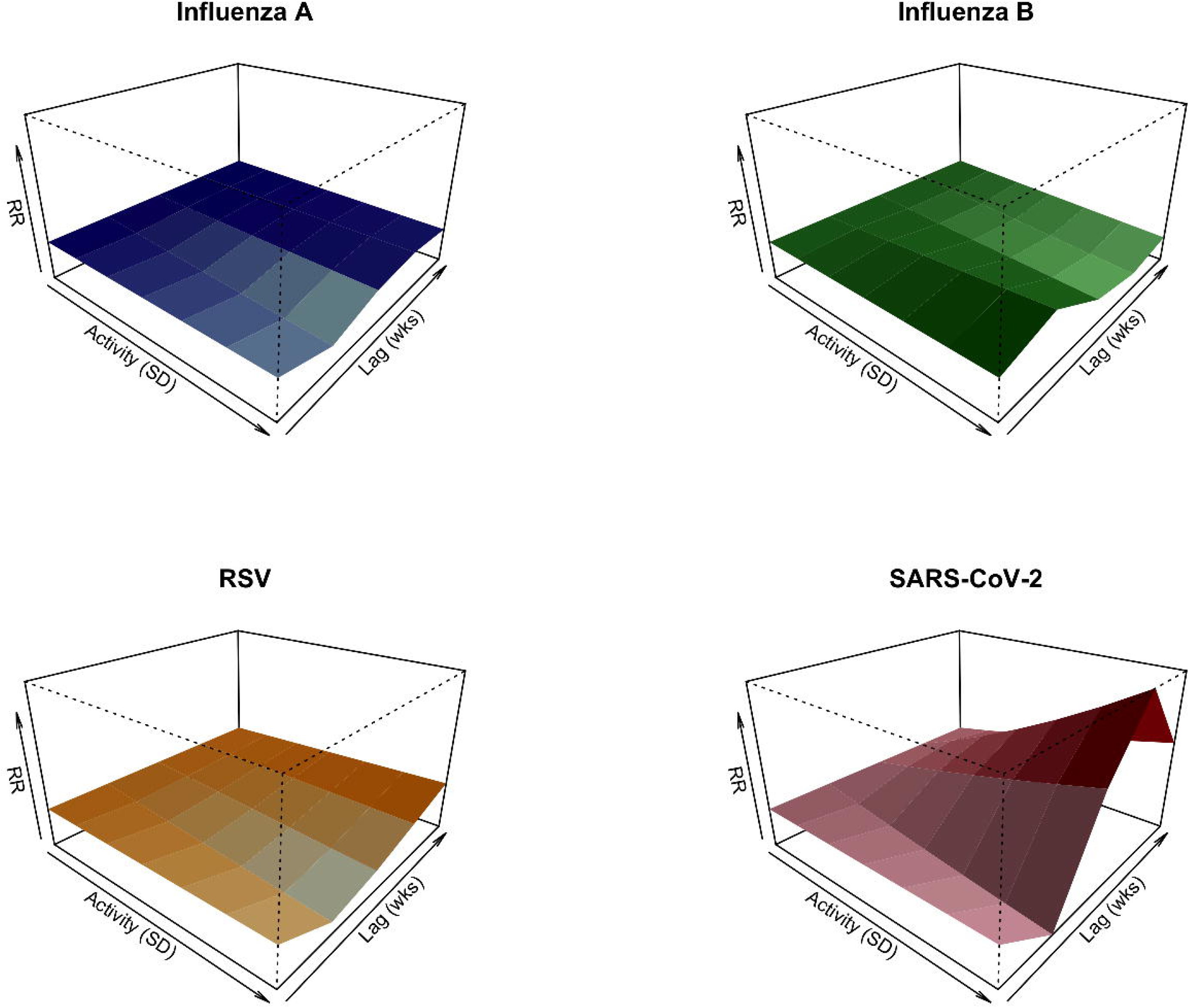
Three-Dimensional Risk Surface Illustrating the Association Between Viral Exposure Intensity, Temporal Lag, and Pulmonary Embolism Risk. The surface plots the non-linear relationship between standardized viral activity (exposure intensity), the time delay following exposure (temporal lag of 1–5 weeks), and the relative risk of pulmonary embolism hospitalization. Darker colors and higher peaks represent periods of elevated risk. (Interactive versions provided as separate digital files).

